# Dynamicity of brain network organization and their community architecture as characterizing features for classification of common mental disorders from the whole-brain connectome

**DOI:** 10.1101/2023.07.14.23292699

**Authors:** Nisha Chetana Sastry, Arpan Banerjee

## Abstract

The urgency of addressing common mental disorders (bipolar disorder, ADHD, and schizophrenia) arises from their significant societal impact. Developing strategies to support psychiatrists is crucial. Previous studies focused on the relationship between these disorders and changes in the resting-state functional connectome’s modularity, often using static functional connectivity (sFC) estimation. However, understanding the dynamic reconfiguration of resting-state brain networks with rich temporal structure is essential for comprehending neural activity and addressing mental health disorders. This study proposes an unsupervised approach combining spatial and temporal characterization of brain networks to classify common mental disorders using fMRI timeseries data from two cohorts (N=408 participants). We employ the weighted stochastic block model to uncover mesoscale community architecture differences, providing insights into neural organization. Our approach overcomes sFC limitations and biases in community detection algorithms by modelling the functional connectome’s temporal dynamics as a landscape, quantifying temporal stability at whole-brain and network levels. Findings reveal individuals with schizophrenia exhibit less assortative community structure and participate in multiple motif classes, indicating less specialized neural organization. Patients with schizophrenia and ADHD demonstrate significantly reduced temporal stability compared to healthy controls. This study offers insights into functional connectivity (FC) patterns’ spatiotemporal organization and their alterations in common mental disorders, highlighting the potential of temporal stability as a biomarker.

## 1. Introduction

Common mental disorders, such as bipolar disorder, Attention-Deficit Hyperactivity Disorder (ADHD), and schizophrenia have a significant societal impact in terms of disability-adjusted life years (DALYs), years lived with disability (YLDs), and years of life lost (YLLs) (Alize J Ferrari, 2022). Developing prevention and intervention strategies to assist behavioural psychiatrists is crucial in addressing these disorders. Neuroimaging techniques like functional magnetic resonance imaging (fMRI) and brain connectivity analytics offer promising non-invasive tools for early detection and management of these disorders (Edgar Canario, 2021) (Miranda, Paul, Putz, Koutsouleris, & Muller-Myhsok, 2021). The spontaneous resting state dynamics of the brain, which is believed to emerge from robust metabolic and neural information processing principles (Deco, Jirsa, & McIntosh, 2010) (Smith, et al., 2013) offers a practical framework for investigating biomarkers of these disorders. However, a systematic and comprehensive approach to pattern detection and biomarker identification is lacking. This study aims to establish a data-driven approach to identify biologically meaningful patterns that differentiate common mental disorders.

Previous studies investigating alterations in resting-state brain dynamics in common mental disorders have largely focused on the strategies of functional connectome communities – the mesoscale organization of individual neural elements into motifs, circuits, or clusters (Betzel, Medgalia, & Bassett, 2018). These studies have reported changes in community organization such as decreased modularity of brain networks in schizophrenia and bipolar disorder (Alexander-Bloch, et al., 2010) (Yu, et al., 2020) (Lerman-Sinkoff & Barch, 2016). Studies using multilayer community detection algorithms have reported higher flexibility in patients with schizophrenia (Gifford, et al., 2020) and ADHD (Ding, et al., 2022). A major drawback of these studies, in addition to the methodological biases of the community detection algorithms (Betzel, Medgalia, & Bassett, 2018) is the utilization of *static* functional connectivity to detect functional connectome communities. However, recent research has shown that resting-state functional brain networks undergo spontaneous, time-varying, and large-scale dynamic reconfiguration (Hutchison, et al., 2013). This suggests that measures assuming stationarity for the entire duration of the scan are too simplistic to capture the full extent of resting brain dynamics (Preti, Bolton, & De ville, 2017).

Dynamic functional connectivity (dFC) can exhibit non-random and non-trivial temporal structures (Lombardo, et al., 2020) and is associated with cognitive processing, learning, attention, and performance (Cohen, 2018), (Bassett, et al., 2011), (Kucyi, Hove, Esterman, Hutchison, & Valera, 2017), (Jia, Hu, & Deshpande, 2014). The dysconnectivity observed in these disorders suggests the presence of complex spatiotemporal alterations in FC (Alexander-Bloch, et al., 2012), (Gifford, et al., 2020). This naturally raises the question: Are there alterations in dFC within the resting state functional connectome in common mental disorders and if so, can they be accurately captured and comprehensively studied? Moreover, can these patterns of dFC serve as characterization tools for common mental disorders? Stability in dFC patterns over time ensures consistent information representation in the neural connectome (Le, Lu, & Yan, 2020). While temporal stability of the brain’s functional connectome has been studied extensively during resting state (Le, Lu, & Yan, 2020) (Sastry, Roy, & Banerjee, 2023), limited research has explored alterations in the temporal stability of the whole-brain dynamic functional connectome in common mental disorders.

To estimate the temporal stability of dFC, researchers often employ methods like K-means clustering (Allen, et al., 2014) (Cabral, et al., 2017) or Hidden Markov models (Viduarre, Smith, & Woolrich, 2017) (Surampudi, et al., 2018) to identify discrete brain states and measure the switching rate or flexibility between these states. Higher flexibility suggests more frequent transitions, which leads to reduced temporal stability (Long, Lu, & Liu, 2023). Graph theoretical studies use *temporal correlation coefficient* to assess dynamic brain network stability. However, these studies are limited in effectiveness due to the ad-hoc selection of the number of states in clustering algorithms (Rakthanmanon, Keogh, Lonardi, & Evans, 2011) (Allen, et al., 2014). An alternative perspective considers dFC as a continuous process, examining global and regional dynamics using techniques like Pearson correlation, angular distance, or dFCSpeed (Zhang, et al., 2016) (Sastry, Roy, & Banerjee, 2023) (Arbabyazd, et al., 2020). This approach captures the continuous fluctuations and patterns in dFC without relying on predefined states.

Thus, the present study has two main objectives. Firstly, it aims to examine the potential of whole brain and sub-network level measures of dFC as characterizing tools for common mental disorders. This investigation is conducted using a large cohort (N=408) comprising healthy individuals as well as those diagnosed with schizophrenia, bipolar disorder, and ADHD. Secondly, the study aims to provide a comprehensive understanding of the impaired brain network mechanisms associated with these disorders by combining community-based detection and dynamics-driven characterization of the functional connectome.

## 2. Material and methods

### 2.1 Participants and Image acquisition

Overall, the study included two multicentre datasets involving 408 human participants.

#### Dataset 1

We downloaded resting state functional magnetic resonance imaging (fMRI) data from 285 participants who participated in the University of California Los Angeles (UCLA) Consortium for Neuropsychiatric Phenomics LA5c study *(Dataset 1)* (Poldrack, et al., 2016) (Gorgolewski, Durnez, & Poldrack, 2017). The public database was obtained via openfMRI (https://openfmri.org/dataset/ds000030/) and includes 138 healthy controls (HC), 58 individuals diagnosed with schizophrenia (SZ), 40 with attention deficit hyperactivity disorder (ADHD) and 49 with bipolar disorder (BP). Further details of the participants, image acquisition and preprocessing of the data are provided in the supplementary material.

#### Dataset 2

For the replication analysis, a publicly available dataset from the centre for Biomedical Research Excellence (COBRE) was obtained (Calhoun, et al., 2012) (Bellec, 2016). The neuroimaging dataset *(Dataset 2)* included resting state functional MRI scans from 72 participants with schizophrenia and 74 healthy controls. Additional details regarding participant characteristics, image acquisition, and data preprocessing can be found in the supplementary material

### 2.2 Data preprocessing and parcellation

The rs-fMRI images were pre-processed using the CONN toolbox (McGovern Institute for Brain Research, MIT, USA) in MATLAB (The MathWorks). The default CONN preprocessing pipeline (defaultMNI) was employed, consisting of functional realignment and unwarp, slice-time correction, outlier identification, direct segmentation and normalization, and functional smoothing. Further details are provided in the supplementary material. The final datasets used in this analysis and their group characteristics are described in *Table S1*. Resting state scans of each participant were parcellated using 400 region Schaefer parcellation (Schaefer, et al., 2018). This atlas was chosen as it pre-allocates brain regions into resting state networks (RSNs). For each subject, mean BOLD time series (*Figure 1A*) were estimated for each region over all voxels belonging to that brain region.

**Figure 1.**
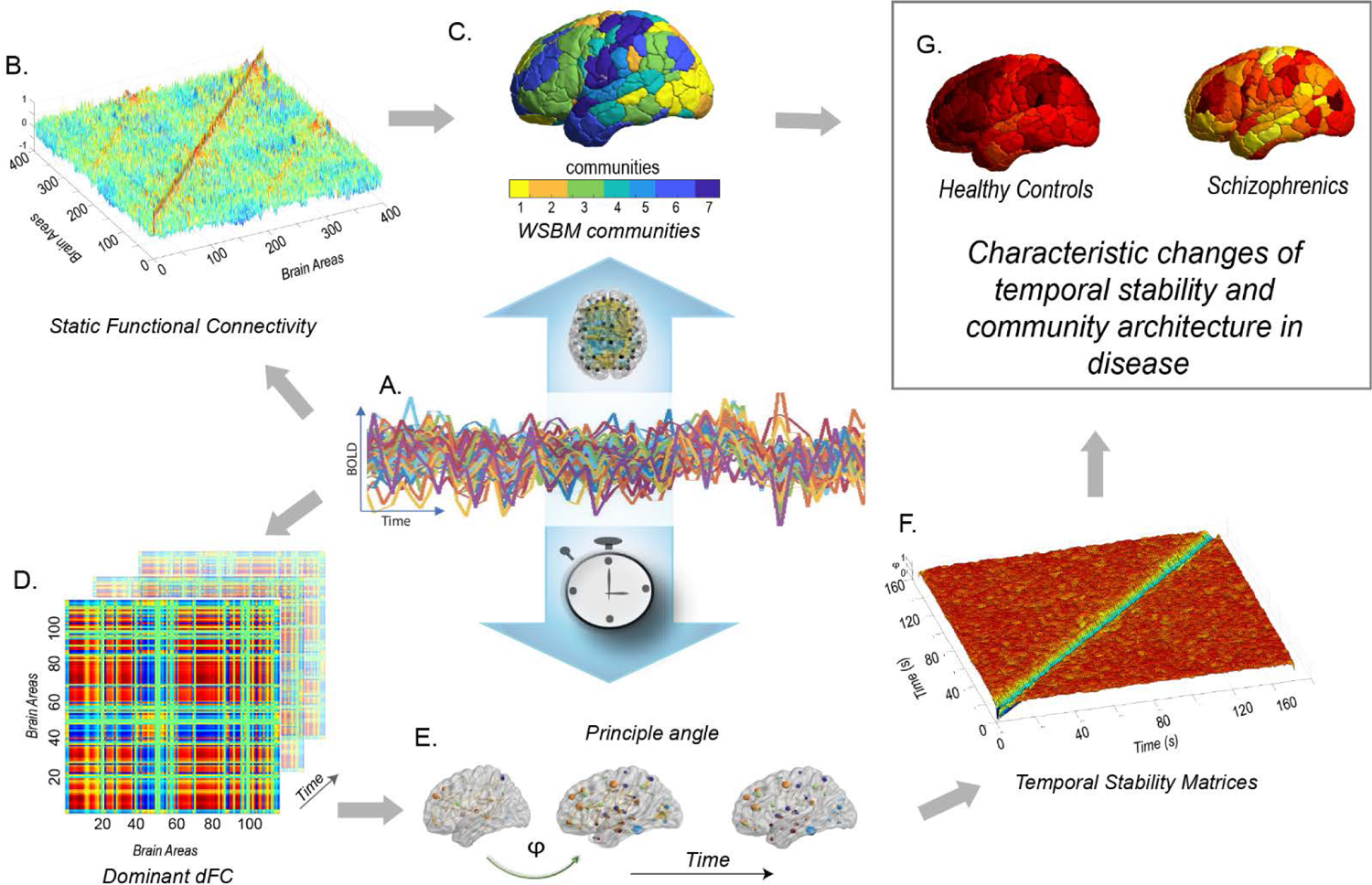
a brief overview of the methodology used in this study. (A) shows the BOLD time series data (B) illustrates the estimation of static functional connectivity using Pearson correlation, which is then transformed using the r-to-z method (C) displays topographic representation of the WSBM communities. The subject-wise undirected, signed, weighted adjacency matrix serves as the input for the Weighted Stochastic Block Model (WSBM), a data-driven generative community detection algorithm that groups brain areas into K=7 communities based on their stochastic equivalence. (D) presents the matrix representation of the reduced dominant dynamic functional connectivity (dFC) patterns, denoted as D(t), computed at each time point (E) demonstrates the calculation of similarity between dominant dFC subspaces using angular distance or principal angle (ϕ) (F) showcases the Time X Time temporal stability matrix, where each entry represents the principal angle (Φ (*t*_x_, *t*_y_) between dominant dFC subspaces at time points *t*_x_ and *t*_y_. This matrix visualizes the temporal landscape, with the principal angle ranging from 0 (indicating low angular distance) to π/2 (indicating high angular distance). Constructing a Time X Time temporal stability matrix allows us to visualize the temporal “landscape” for the entire duration of the scan. (G) we calculate the temporal stability of the dynamic functional connectome using two measures: entropy and global temporal distance. We investigate changes in community architecture in common mental health disorders such as schizophrenia, bipolar disorder, and ADHD.

### 2.3 dominant dFC and temporal stability matrices

These methods have first been introduced in (Sastry, Roy, & Banerjee, 2023) in the context of healthy ageing. For each subject, we estimate time-resolved dFC using BOLD phase coherence (*Figure 1A*) (Glerean, Salmi, Lahnakoski, Jaaskelainen, & Sams, 2012) (Cabral, et al., 2017), which resulted in a matrix with size N X N X T, where N = 400 is the number of brain regions, T is the total number of time points (T = 152 for *Dataset 1*, and T= 150 for *Dataset 2*). First, instantaneous phases *θ (n, t)* of the BOLD signals for all the brain regions was calculated using Hilbert transform. Given the phases of BOLD signals, phase coherence between brain areas *n* and *p* at each timepoint *t*, i.e., *dFC (n, p, t)* is computed as:

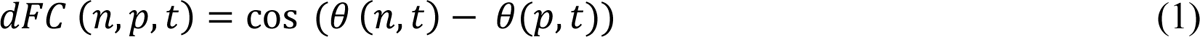

To characterize the evolution of dFC over time, we extract dominant subspace of dFC patterns (*Figure 1B*), by applying PCA (Friston, Frith, Liddle, & Frackowiak, 1993) (Sastry, Roy, & Banerjee, 2023). Given a set of dominant dFC matrices (D_*t*_), we seek to characterize temporal stability using the similarity of dFC patterns across timepoints. We use angular distance (*Figure 1C*) to estimate the similarity between dominant dFC subspaces. A detailed overview of the methodology can be found in (Sastry, Roy, & Banerjee, 2023).

### 2.4 Measures of temporal stability

Constructing a *time X time* temporal stability matrix allows us to visualize the ‘*temporal landscape’* (*Figure 1D*) for the entire duration of the scan. We introduce two distinct perspectives on measures of temporal stability – Firstly, we seek to quantify temporal stability over the entire temporal landscape across all timepoints. To achieve this, we evaluate the informational content of the stability matrices by calculating entropy (Sastry, Roy, & Banerjee, 2023). Entropy was used because it is a more direct measure of order and disorder in a dynamical system and provides us a measure of distinguishable temporal order that can be interpreted as the overall stability of the temporal landscape (Yang, et al., 2013). Entropy is defined by the following equation:

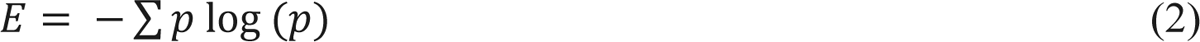

Where *p* contains normalized histogram counts returned from “imhist.m” applied on temporal stability matrices, estimated using reduced D_*t*x_ and D_*t*y_. “imhist.m” calculates the histogram of temporal stability matrices and returns the normalized counts. Overall temporal stability is estimated as follows

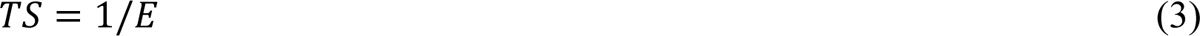

Secondly, we seek to quantify temporal stability across successive time windows. To do this, we estimate the *global temporal distance* by taking the average of the off-diagonal elements of the temporal stability matrix. The off-diagonal elements are angular distances among dominant dFC subspaces at two successive time points, thus the term ‘*global’* signifies that the measure captures a temporally averaged snapshot of the dFC evolution.

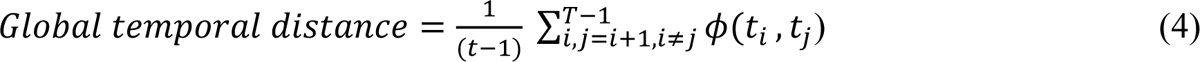

Where, Φ (*t*_i,_ *t*_j_) is the angular distance entry at *i^th^* row and *j^th^* column in the temporal stability matrix, T is the total number of timepoints. *Global temporal stability (TSglobal)* is defined as the inverse of the *global temporal distance*:

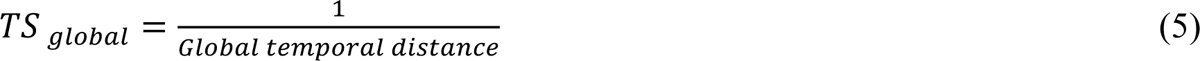

### 2.5 Model-based community detection using the weighted stochastic block model (WSBM) on static FC

We estimate the sFC (Friston, 2011) between two brain areas, *n, p* by calculating the Pearson correlation between BOLD time series of brain areas *n, p* (Biswal, Yetkin, Haughton, & Hyde, 1995) (*Figure 1E*). The correlations were subsequently r-to-z transformed. The *N X N* FC matrix was represented as a network in which regions were represented by network nodes and FC between region *n* and *p* was represented by network edge between nodes *n* and *p* (Bassett, Zurn, & Gold, 2018). Thereafter, subject-wise undirected, signed, weighted adjacency matrix *(A)* was estimated for detection of community architecture. For each subject, for a given *N* x *N* adjacency matrix, we estimate WSBM and maximize the likelihood using variational bayes algorithm described by (Aicher, Jacobs, & Clauset, 2015). We select *k=*7 (number of communities) (Tooley, Bassett, & Mackey, 2022) (Allen, et al., 2014) and repeat the optimization procedure 30 times for each subject. We implement the WSBM procedure in MATLAB using freely available code and estimate a WSBM for each subject (https://aaronclauset.github.io/wsbm/) (Tooley, Bassett, & Mackey, 2022). Additional details on WSBM can be found in supplementary material.

### 2.6 Measures for community architecture and interaction motifs

WSBM assigns brain areas into communities (*Figure 1F*). We characterize the interaction between communities using interaction motifs described in (Betzel, Medgalia, & Bassett, 2018). One dimension on which we characterized the community interaction was the extent to which detected communities were assortative. The interaction between two communities, *r,* and *s,* can be characterized by the community densities:

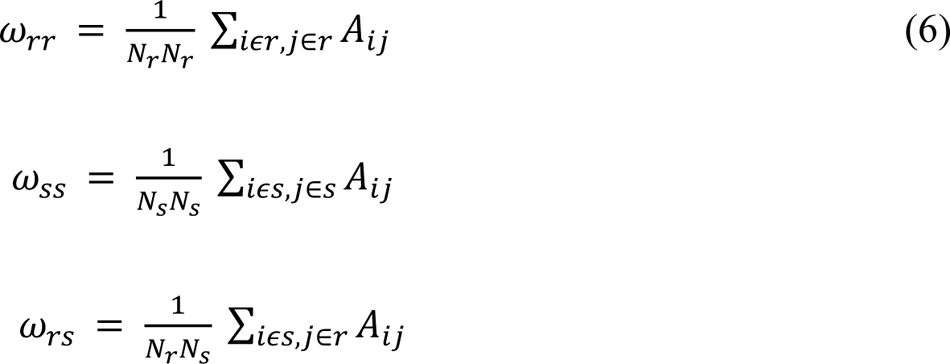

Where N_*r*_ and N_s_ are the number of nodes assigned to communities *r* and *s* and *A* is the adjacency matrix. Given these community densities, we classify their interactions as follows:

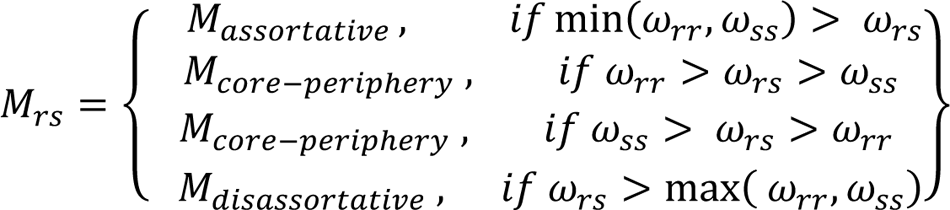

Although interaction motifs are defined at the level of communities, the motifs can be mapped and an analogous score for individual brain regions can be calculated. Given a region *i’s* community assignment Z_i_, its connection density to a community *r* is given by

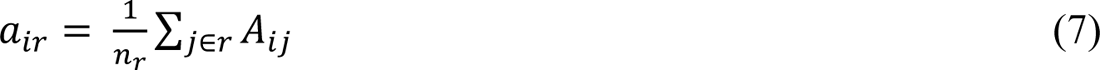

Then the regional assortativity score is given by:

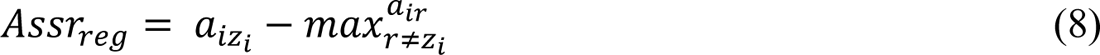

While calculating both regional and community assortativity scores, singleton communities have been excluded (Betzel, Medgalia, & Bassett, 2018).

### 2.7 Diversity Index

In addition to studying interaction motif classes (assortative, core, periphery), for each motif class, we calculate how frequently it appears among community *r’*s interactions. For a *k* community partition, community *r* participates in *k – 1* interaction. The frequencies of appearance can be expressed as probabilities, P_a_, P_c,_ P_*p*_, and P_d_(“assortative”, “core”, “periphery”) and we can then calculate the entropy as:

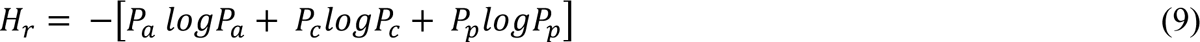

The entropy or Diversity Index is *0* if the community (*r*) participates in only one class and is maximized if *r* participates in all classes equally (Betzel, Bertolero, & Bassett, 2018). The resulting score is then assigned to all the nodes cc ∈ *tt*. We calculate this for all *k* communities and estimate mean diversity index by averaging across communities.

### 2.8 Morphospace analysis

We adopt this analysis from (Betzel, Medgalia, & Bassett, 2018). A morphospace is a hyperspace whose axes represent the features of the organism or a system. Network morphospace represents the topological properties of a network and helps visualize the richness of the topology (McGhee, 2006). In this study, we construct a community morphospace, whose axes are within-community (ω_*rr*,_ ω_ss_) and between-community densities (ω_*rs*_). Each point in the morphospace represents a pair of communities, *r,* and *s*.

## 3. Results

We report on how the community architecture and temporal characteristics of functionally connected brain networks can be used as a characterization tool for common mental disorders. The community architecture was assessed using the WSBM to uncover the meso-scale community structure from sFC (see methods section). We used three network topology measures that indicate each brain area’s participation in assortative, core, or peripheral community interactions (Betzel, Bertolero, & Bassett, 2018) and demonstrate their distinctness across common mental disorders. We examined temporal properties using the temporal stability of functionally relevant brain networks, employing established methods (Sastry, Roy, & Banerjee, 2023). This involved capturing temporal fluctuations in dFC, constructing a *temporal landscape* with temporal stability matrices. We computed entropy and global temporal distance metrics to quantify the informational content within these matrices, serving as defining characteristics for mental health disorders such as schizophrenia, ADHD, and bipolar disorder. Section 1 presents community architecture analysis results and Section 2 focuses on temporal stability analysis for *dataset 1* (Poldrack, et al., 2016) in both diseased and healthy cohort. To ensure validity, we replicate the pipeline with *dataset 2* (Bellec, 2016), specifically examining participants with schizophrenia in Section 3.

### 3.1 Distribution of community architecture of brain networks across spectrum of common mental disorders

In *dataset 1*, a WSBM (see Methods for details) was fitted on the adjacency matrix for each subject in the diseased and healthy cohorts. The number of communities was set at k=7 based on previous studies (Allen, et al., 2014) (Tooley, Bassett, & Mackey, 2022). *Figure 2A* shows the community partitions in FC for both diseased and healthy individuals, while *Figure 2B* presents a topographic representation of the communities detected with WSBM in individuals with schizophrenia, ADHD, bipolar disorder, and healthy controls. Distinct assignments of brain areas to different communities were observed in diseased and healthy controls (*Figure 2B*). Community motifs reflect assortative, core, or peripheral interactions among pairs of communities. In assortative communities, the internal density of connections within subnetworks exceeds their external density whereas core-periphery organization consists of a central core which is connected to the rest of the subnetworks and peripheral nodes connect to the core but not with each other (Betzel, Bertolero, & Bassett, 2018). Here in the main manuscript, we report only significant group-level community interaction measures averaged across communities. *Figure 2C* reports significantly lower assortativity in schizophrenics *(P=0.0121, t=2.5374)* compared to healthy controls. The distributions were parametric and unpaired t-test was used to assess significance. Rank sum test (the distribution was non-parametric) revealed a significantly higher coreness in schizophrenics *(P=0.0188)* compared to healthy counterparts. Additionally, the motif participation index and diversity index were computed for individual brain areas. *Figure 2C* reveals a significantly higher diversity index in schizophrenics *(unpaired t-test, P=0.0354, t = −2.1207)* indicating their communities, by and large, participate in more than one motif class. Although distinct modifications in community interaction motifs were observed for individuals with bipolar disorder and ADHD compared to healthy controls (see supplementary *S 1*) the results were non-significant. Next, a 3D community morphospace was constructed, where each point represents a pair of communities *{r,s}*, and the axes are defined by within-community and between-community densities, ω_*t*_, ω_*t*c_, ω_c_. Morphospace analysis revealed that individuals with schizophrenia, ADHD, and bipolar disorder favoured fewer assortative and included more core-periphery community interactions than healthy controls *(Figure 2D)*. Overall, the community motifs and morphospace analysis indicate community structure in common mental disorders, especially in schizophrenics is less assortative.

**Figure 2.**
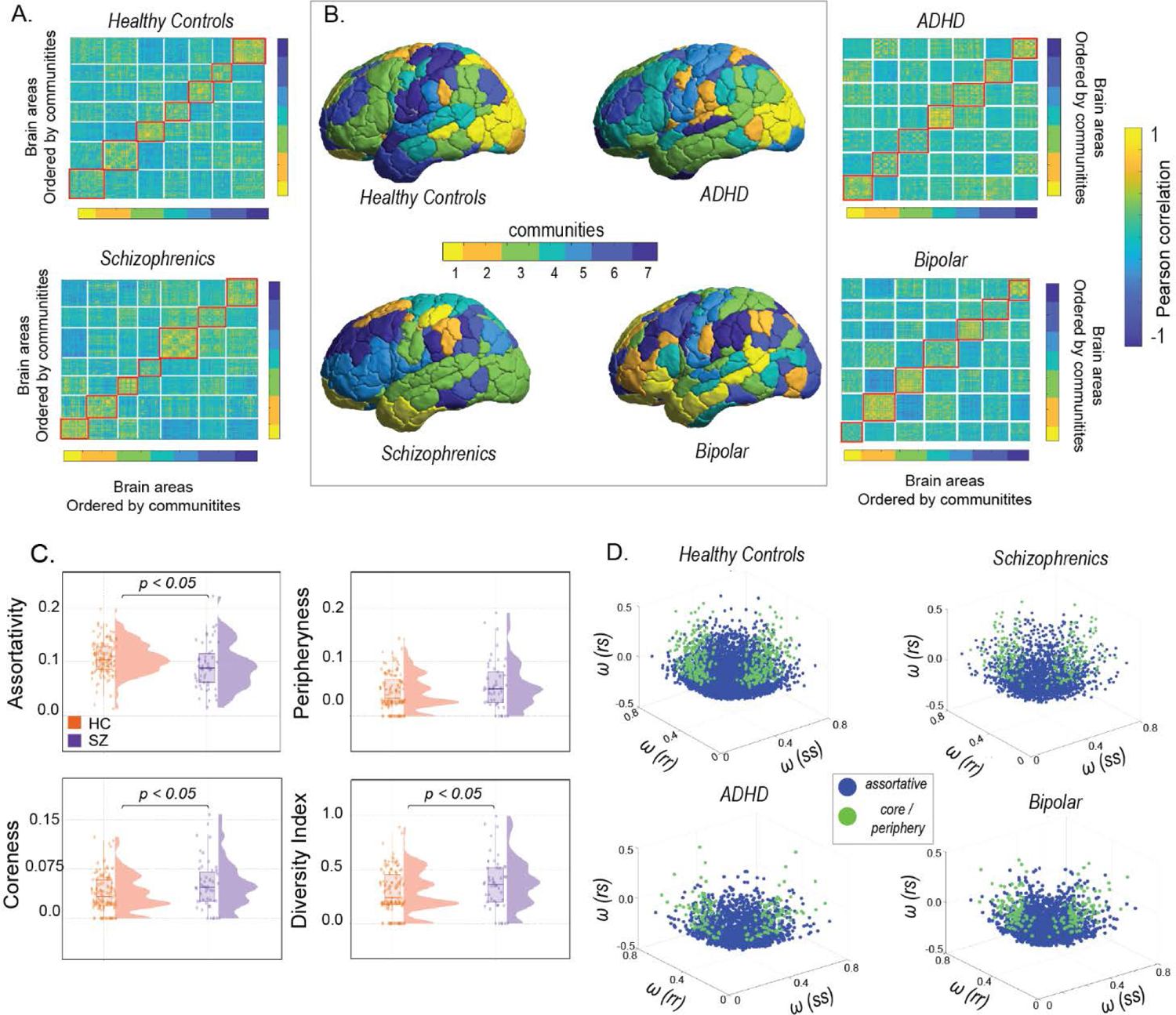
An overview of the community architecture differences between schizophrenics and healthy controls in Dataset 1. (A) shows the static functional connectivity matrices of schizophrenics and healthy controls, with brain areas ordered by communities (B) presents the partition of cortical regions into 7 communities using the Weighted Stochastic Block Model (WSBM), a generative community detection algorithm that groups stochastically equivalent brain regions into communities (C) each pair of communities (r and s) is classified into one of three community motifs: assortative, coreness, and peripheryness. The diversity index is calculated as the average across all brain regions per subject. The violin plots indicate that in schizophrenics, communities are less assortative (D) illustrates the construction of a network morphospace using all pairs of communities, which are coloured according to their motif type: blue for assortative community interactions and green for core or periphery community interactions.

Next, to identify disease-specific changes in brain regions, an analogous assortativity score was calculated for each region in patients with schizophrenia, ADHD, and bipolar disorder from *dataset 1* (see methods) (Betzel, Medgalia, & Bassett, 2018). Age-matched and sex-matched healthy controls were generated for each disease group. For each brain region, a t-test was performed between the assortativity scores of patients and matched healthy controls, with age and sex regressed out. In patients with schizophrenia, significant decreases in assortativity were observed in the peripheral visual, Dorsal attention, Ventral attention, and Temporal parietal networks (*Table S2*) whereas for individuals with ADHD, significant decreases in assortativity were found in the central visual, limbic, dorsal attention, default, and control networks (*Table S4*). In individuals with bipolar disorder, significant reductions in assortativity were seen in the dorsal and ventral attention, limbic, and default networks (*Table S3*). *Figure 3A-C* shows brain-wide topography of significant assortativity scores between patients with ADHD, bipolar disorder, schizophrenia and healthy controls.

**Figure 3.**
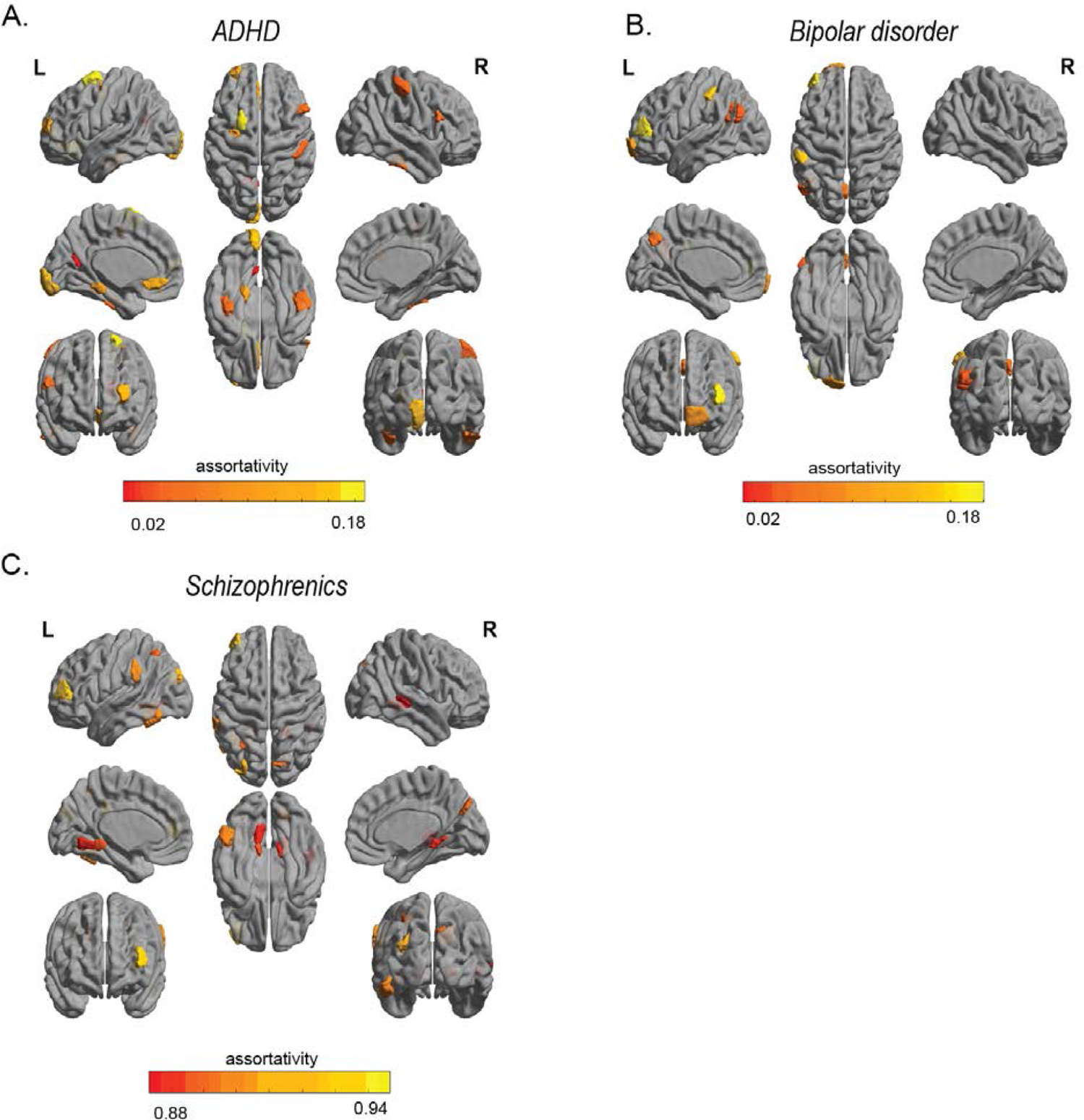
Profile of brain regions showing significant difference in assortativity between patients, age and sex matched healthy controls. (A) ADHD (B) bipolar disorder (C) Schizophrenia.

### 3.2 Impaired and preserved patterns of temporal stability in common mental disorders: schizophrenia, ADHD and bipolar

To compute the temporal stability of dFC, the first step involved estimating the similarity/differences between dominant dFC subspaces by calculating the angular distance between them (Φ (*t*_x_, *t*_y_(). The resulting temporal stability matrix, spanning time X time, characterizes the collective temporal characteristics of dFC and aids in visualizing the *temporal landscape*. A low angular distance between subspaces indicates that the corresponding dFC patterns were similar in configuration, while a high angular distance suggests dissimilarity (Sastry, Roy, & Banerjee, 2023). In this section, we present the results of evaluating temporal stability changes between healthy controls and cohorts with common mental disorders (such as schizophrenia, ADHD, and bipolar disorder) in *Dataset 1*.

Group-level averages of temporal stability, computed on resting-state fMRI BOLD time series from healthy controls and individuals with schizophrenia, are shown in *Figure 4A*. Schizophrenics exhibit shorter-lived, low angular distance (yellow hue) repeated patterns of stability, while healthy controls display a more evenly distributed pattern. Quantifying the differences, we calculated the entropy of temporal stability matrices *Figure 4B*, revealing higher entropy in schizophrenics, indicating impaired temporal stability. The distributions were assessed parametrically using the Jarque-Bara test and D’Agostino-Pearson omnibus test. A two-sample t-test revealed significant differences in entropy values between healthy controls and individuals with schizophrenia (P=0.0475, t=-1.9963). Global temporal distance (see Methods for details) also suggests higher values in individuals with schizophrenia. Although the results (*Figure 4C*) were not statistically significant (Wilcoxon rank-sum test, P=0.8902), the violin plots suggest a trend: individuals with schizophrenia exhibit higher global temporal distance compared to healthy controls. Analysing the 8 resting-state networks (Schaefer, et al., 2018) using the same pipeline, we observed significantly higher global temporal distance (*Figure 4D*) in the Dorsal attention network (P=0.0153, t=-2.4489) (The distributions were assessed parametrically using an unpaired two-sample t-test) and Somatomotor network (P=0.0109) (The distributions were non-Gaussian; hence, the Wilcoxon rank-sum test was used), indicating significantly lower temporal stability in individuals with schizophrenia. Overall, these results indicate impaired temporal stability in individuals with schizophrenia, both at the whole-brain level and within specific networks.

**Figure 4.**
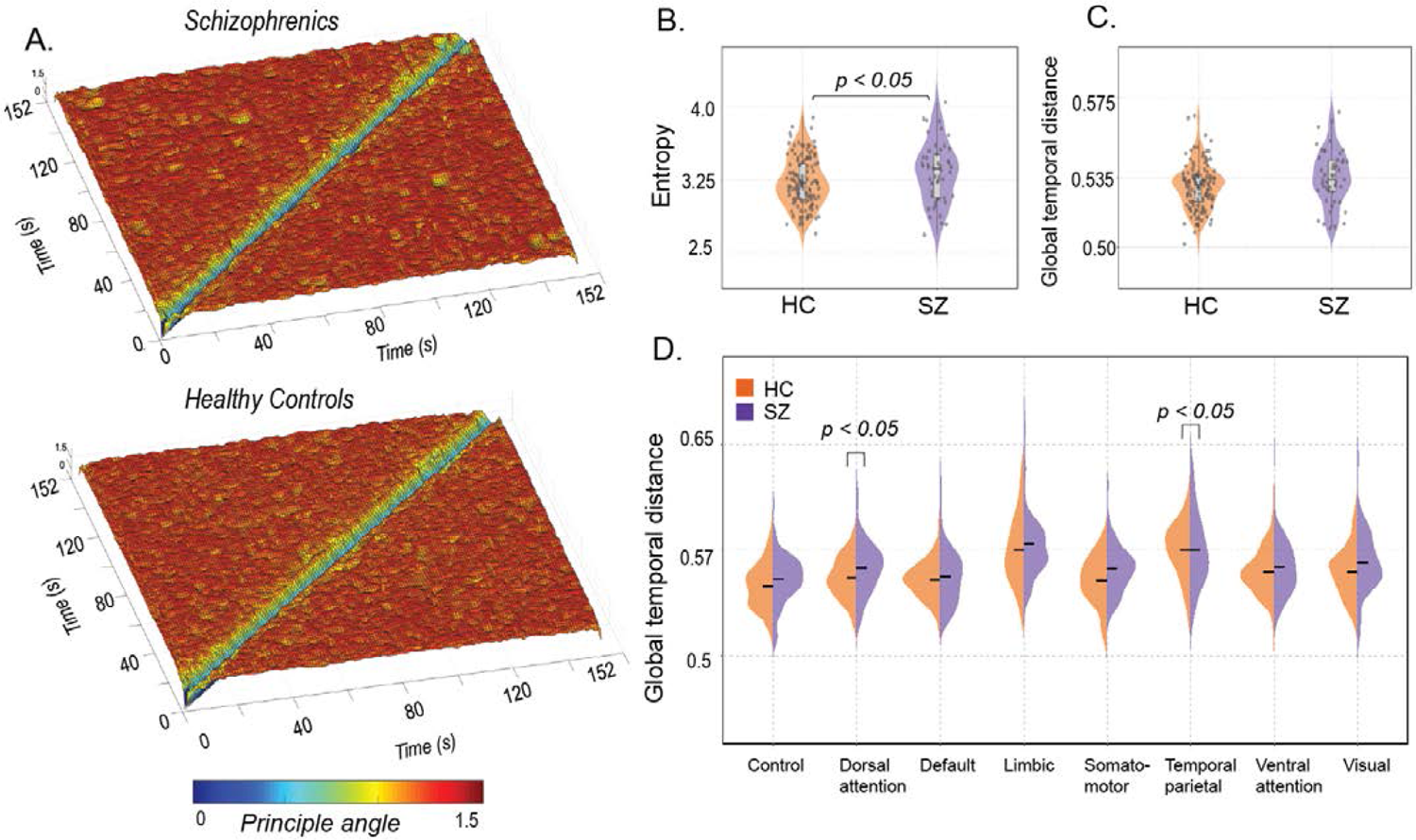
An overview of the temporal stability differences between schizophrenia and healthy controls in Dataset 1. (A) showcases the Time X Time temporal stability matrices, visualized as the *’Temporal landscape’.* Each entry represents the angular distance between dominant dFC subspaces at time points *t*_x_ and *t*_y_, for patients with schizophrenia and healthy controls. In schizophrenics, there is a global spread of shorter-lived, low angular distant (yellow hue) repeated patterns of stability (B) focuses on quantifying temporal stability across the entire Time X Time temporal landscape using entropy (C) quantifies temporal stability over successive time points using global temporal distance. Both measures indicate low temporal stability in schizophrenics (D) global temporal distance is estimated for all the resting state networks defined in the Schaeffer atlas, comparing schizophrenics (purple) and healthy controls (orange).

We further examined temporal stability in patients with other common mental disorders, namely ADHD, Bipolar disorder, and healthy controls from *Dataset 1.* Whole-brain temporal stability matrices were constructed for each participant, revealing minimal spread of low angular distant (yellow hue) repeated stability patterns in both ADHD and Bipolar disorder. Participants with ADHD showed significantly lower entropy (two-sample t-test, P=0.0162, t=-2.4293), and higher global temporal distance (Wilcoxon rank-sum test, p=2.2128e-21), indicating impaired temporal stability (*Figure 5B*). In contrast, participants with bipolar disorder displayed higher entropy (two-sample t-test, P=0.3467, t=0.9436) and global temporal distance (two-sample t-test, P=0.8892, t=0.1395) of temporal stability matrices, although these differences were not statistically significant.

**Figure 5.**
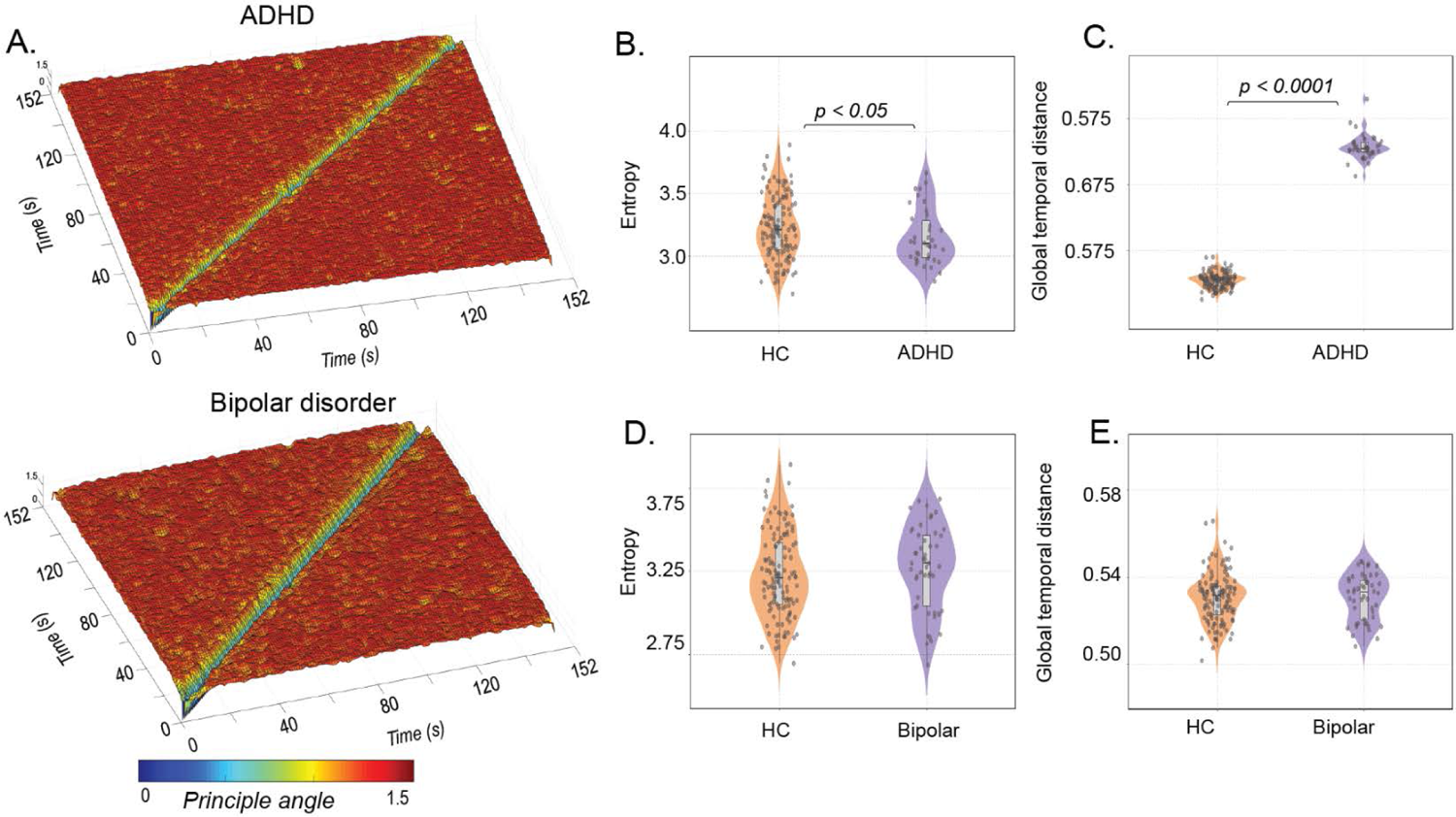
An overview of the temporal stability differences in participants with ADHD and Bipolar disorder. (A) showcases the Time X Time temporal stability matrices, where each entry represents the angular distance between dominant dFC subspaces at time points *t*_x_ and *t*_y_, for participants with ADHD and Bipolar disorder. The temporal stability is quantified using two measures: entropy (B) and global temporal distance (C) in patients with ADHD and healthy controls. The results show that global temporal distance is significantly higher in participants with ADHD, indicating decreased temporal stability. Similarly, temporal stability is quantified using entropy (D) and global temporal distance € in participants with Bipolar disorder and healthy controls.

### 3.3 Validation Analysis

Furthermore, we replicated the entire analysis pipeline using *Dataset 2*. Community detection using the WSBM revealed significant differences in community interaction motifs between individuals with schizophrenia and healthy controls (see Supplementary*, S 2 A*). Schizophrenia patients exhibited lower assortativity and higher coreness and peripheryness (Wilcoxon rank-sum test; P=0.0483, P=0.0209, P=0.0043, respectively). The diversity index was also significantly higher in schizophrenia patients (*S 2 B*) (Wilcoxon rank-sum test, P=0.0184), indicating participation in multiple motif classes. Additionally, the 3D community morphospace analysis validated these findings, specifically in patients with schizophrenia, showing fewer assortative interactions in individuals with schizophrenia (*S 2 C*). Temporal stability analysis confirmed impaired temporal stability in schizophrenia (*Figure 6A*) patients, with higher entropy (Wilcoxon rank sum test, p=0.0373) (*Figure 6B*) and global temporal distance (Wilcoxon rank-sum test; p=2.5559e-04) (*Figure 6C*). At the network level analysis (*Figure 6D*), we observe significantly higher global temporal distance in individuals with schizophrenia in the Control network (p=5.5055e-04), Dorsal attention network (p=0.0057), Default network (p=0.0048), Somatomotor network (p=8.021e-04), Temporal-Parietal network (p=0.0106), and Ventral attention network (p=0.0042). The distributions were non-parametric, and significance was tested with the Wilcoxon rank-sum test. These results validate our initial findings from *dataset 1*, supporting the impaired temporal stability in schizophrenia.

**Figure 6.**
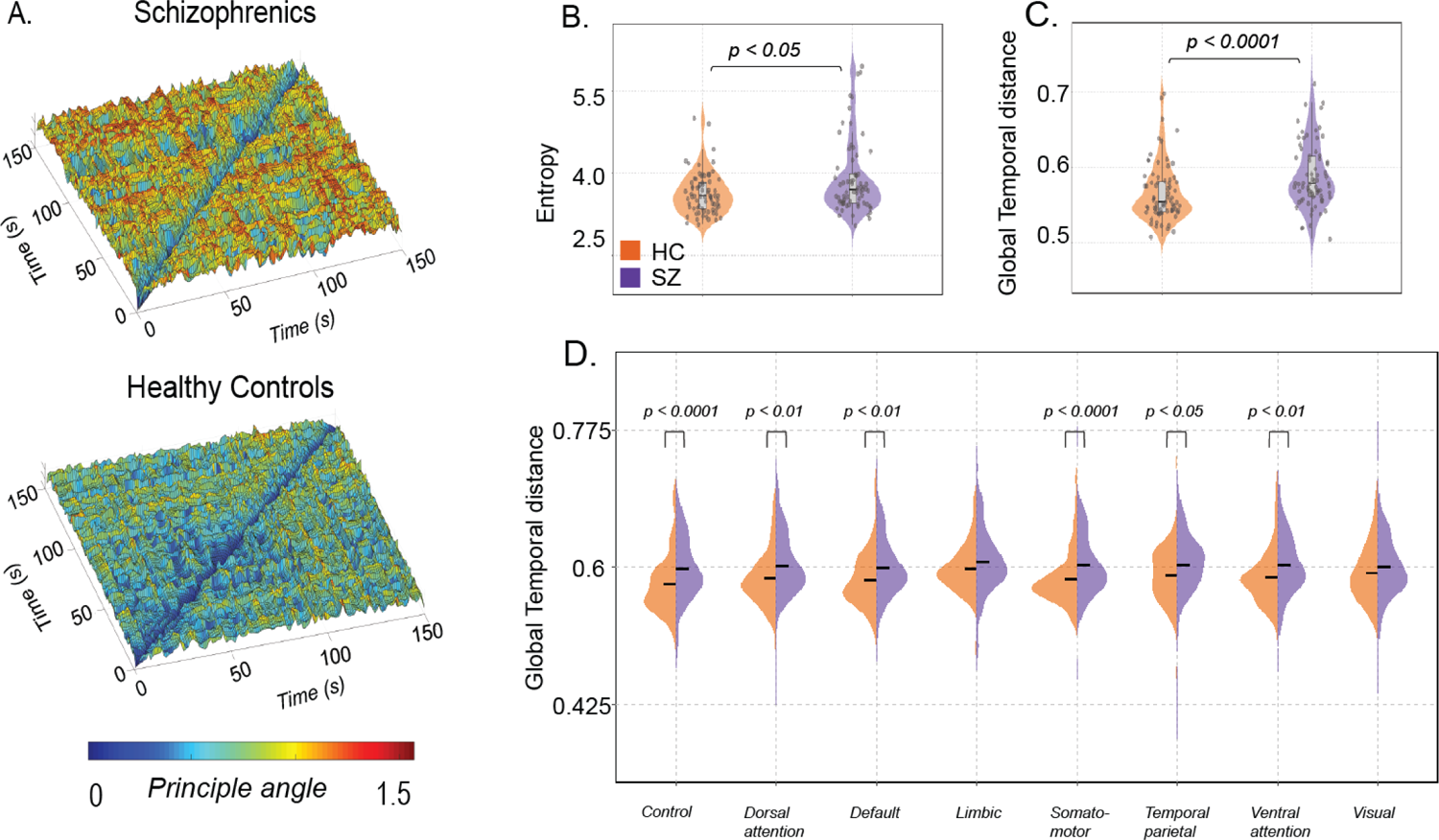
An overview of the temporal stability differences between individuals with schizophrenia and healthy controls in Dataset 2. (A) illustrates the temporal stability matrices, visualized as a temporal landscape, for both schizophrenics and healthy controls. Each entry in the matrix represents the angular distance between dominant dFC subspaces at time points *t*_x_ and *t*_y_. The angular distance ranges from 0 (indicated by yellow hue) to π/2 (indicated by red hue) (B) quantifies the temporal stability across the entire temporal landscape using entropy (C) quantifies the temporal stability over successive time windows using global temporal distance. Both measures indicate a decrease in temporal stability in schizophrenics, which aligns with the findings from Dataset 1. (D) estimates the global temporal distance for each of the resting state networks defined in the Schaeffer atlas for both individuals with schizophrenia (purple) and healthy controls (orange). The plots demonstrate a significant decrease in temporal stability in schizophrenics, particularly in the control, dorsal attention, default, somatomotor, temporal parietal, and ventral attention networks.

## 4. Discussion

The study aimed to explore dFC measures as tools for characterizing common mental disorders disorder. Using two datasets (N = 408), the WSBM (Aicher, Jacobs, & Clauset, 2015) revealed lower assortativity and participation in multiple motifs in schizophrenia compared to healthy individuals (*Figure 2* and *supplementary S 2*). Comparing across diseases with matched controls, specific brain areas showed significant differences in assortativity (*Figure 3*). Although altered community interactions were observed in ADHD and bipolar disorder, statistical significance was not reached (see supplementary *S 1*). To address this limitation, alterations in the dFC were investigated using a novel methodology (Sastry, Roy, & Banerjee, 2023). This approach leverages FC dynamics to assess temporal stability in participants with ADHD, bipolar disorder, schizophrenia, and healthy controls. Entropy and global temporal distance were used as measures of temporal stability. Results showed increased entropy in the whole-brain temporal landscape and specific resting-state networks in schizophrenia, indicating decreased temporal stability of dFC (*Figure 4*). ADHD participants exhibited a significant decrease in temporal stability (*Figure 5*). Notably, temporal stability and community architecture alternations in schizophrenia were consistent across the two datasets used (*Figure 6*).

Prior studies have shown altered community structure in mental disorders, particularly in schizophrenia. Modularity maximization studies reported decreased modularity in functional brain networks of individuals with schizophrenia (Alexander-Bloch, et al., 2010). Other studies using normalized mutual information found significant differences in community structure, with subcortical, auditory, and somatosensory networks being key contributors (Lerman-Sinkoff & Barch, 2016). However, these studies were limited by biases associated with modularity maximization and infomap techniques (Betzel, Medgalia, & Bassett, 2018). To overcome these limitations, the WSBM, capable of uncovering assortative and non-assortative communities was used in this study (Betzel, Medgalia, & Bassett, 2018). WSBM revealed distinct communities in diseased and healthy controls (*Figure 2B* and *S 2 A).* In our analysis, we classified community interactions into assortative, core, and periphery motif classes. Results showed less assortativity, higher coreness and peripheryness in communities of individuals with mental disorders, particularly schizophrenia, compared to healthy controls (*Figure 2C, Figure 2D* and *S 2*). The Diversity Index indicated greater participation in multiple motif classes in schizophrenia (*Figure 2C* and *S 2 B*). Furthermore, significant changes in assortativity at the regional level were observed in schizophrenia, bipolar disorder, and ADHD. These findings provide insights into the neuropathological profiles of these common mental disorders.

Previous studies in schizophrenia have identified disruptions in modular structure in sensory, auditory, and visual areas (Bordier, Nicolini, Forcellini, & Bifone, 2018) (Alexander-Bloch, et al., 2012). In our study, we found significantly lower assortativity in peripheral visual, dorsal and ventral attention, and temporal parietal networks in schizophrenia (*Figure 3C) (Table S2)*. ADHD patients have shown decreased brain network integration and increased network segregation, with significant alterations in local clustering coefficients in cerebellar, frontal, motor, and temporal regions (Lin, et al., 2014). We observed significant alterations in assortativity in central visual, limbic, default, and somatomotor networks in ADHD patients (*Figure 6*A) (*Table S4*). Studies on bipolar disorder have identified alterations in brain network topology in frontoparietal and limbic networks (Zhang, et al., 2021). Similarly, our findings indicate significantly lower assortativity in default and dorsal attention networks, as well as limbic networks, in bipolar disorder patients (*Figure 3*B) (*Table S3*). Overall, these region-level alterations align with previous studies and suggest that alterations in community structure play a role in the neuropathology of common mental disorders. Although our attempts to replicate community detection and interaction motif analysis in ADHD and bipolar disorder patients did not yield statistically significant results, we attribute this to the limited number of participants and the dependence of most community detection algorithms on sFC. It is crucial to acknowledge the limitations of sFC in comprehensively capturing the dynamic nature of brain activity (Preti, Bolton, & De ville, 2017). Therefore, we focused on estimating the temporal stability of the resting-state dynamic functional connectome to explore its dynamic characteristics and alterations in common mental disorders. Our methodology involved extracting dominant dFC patterns at each time point, projecting them into dFC subspaces using PCA, and assessing their similarity through angular distance calculations (Sastry, Roy, & Banerjee, 2023) (*Figure 1C*). We introduce two measures to quantify temporal stability: 1) Entropy, which estimates temporal stability across the entire temporal landscape and all time points, and 2) Global temporal distance, which measures temporal stability across successive time points. While previous studies have explored regional variations in temporal stability (Zhang, et al., 2016) (Dong, et al., 2019), our study extends this exploration by quantifying temporal stability at both the whole-brain and brain network level. Our key finding is a significant decrease in temporal stability among individuals with schizophrenia at both the whole-brain and network levels. In both *Dataset 1* and *Dataset 2,* we observed increased entropy (*Figure 4B*) (*Figure 6B*) and global temporal distance (*Figure 4C*) (*Figure 6C*) of the whole-brain functional connectome in individuals with schizophrenia, indicating impaired temporal stability as a marker of the disease. Furthermore, several sub-networks, including the dorsal attention, somatomotor, limbic, ventral attention, control, and temporal-parietal networks, exhibited impaired temporal stability (*Figure 4D* and *Figure 6D*). One earlier study (Zhang, et al., 2016) found that patients with schizophrenia exhibited significantly increased temporal variability (decreased temporal stability) of dFCs in subcortical regions, such as the thalamus, palladium, and visual areas during resting state. Similarly, using flexibility measures, few groups (Dong, et al., 2019), (Gifford, et al., 2020) (Long, et al., 2020), have observed significant impairment in temporal stabilities of dFC in multiple brain areas, including the thalamus, visual areas among individuals with schizophrenia. In line with these previous studies, our results emphasizing the significant impairment in temporal stability of dFCs at both the whole-brain and network level have contributed towards a comprehensive understanding of the temporal stability differences observed in schizophrenia. ADHD is characterized by dynamic reconfiguration of the functional connectome, with increased temporal variability in the default-mode network and lower temporal variability in subcortical regions reported in previous studies (Lin, et al., 2014) (Fair, et al., 2010) (Zhang, et al., 2016) (Castellanos, et al., 2008). Our findings show a significant increase in the global temporal distance of the whole-brain functional connectome in ADHD patients, indicating decreased temporal stability (*Figure 5B)*. Similarly, there are multiple studies reporting shared similarities in temporal stability alterations between bipolar disorder and schizophrenia (Long, Lu, & Liu, 2023) (Han, et al., 2020) (Nguyen, et al., 2017). Although our results show changes in temporal stability in individuals with bipolar disorder, the results were not statistically significant. Overall, our findings, validated across two datasets, highlight a significant widespread decrease in the temporal stability of dFC in common mental disorders, especially in schizophrenia. These alterations in dynamic functional network configurations, captured by temporal stability measures, provide a comprehensive representation of brain dynamics and reflect the dynamic nature of mental disorders. They hold promise as potential biomarkers for common mental health pathologies, particularly in ADHD and schizophrenia, where mental state fluctuations are more dynamic compared to bipolar disorder.

A major methodological limitation of our study is with WSBM. WSBM requires the user to specify the number of communities (K). Based on previous studies, we chose K=7, but this choice may impact the results (Tooley, Bassett, & Mackey, 2022) (Allen, et al., 2014). In conclusion, our study emphasizes the importance of dFC and temporal stability in characterizing common mental disorders. While WSBM reveals distinct patterns in the community architecture of functionally connected brain networks between diseased and healthy controls, it faces challenges in differentiating between specific disorders. However, temporal stability analysis using angular distance, global temporal distance, and entropy calculations uncovers impaired stability in schizophrenia and ADHD, while bipolar disorder exhibits notable differences. These findings highlight the significant role of dFC and temporal stability in understanding and characterizing common mental disorders, providing insights into underlying mechanisms and potential diagnostic markers.

## Ethics statement

The COBRE (The Centre for Biomedical Research Excellence) dataset was obtained through the International Neuroimaging Data-sharing initiative (http://fcon_1000.projects.nitrc.org/indi/retro/cobre.html). This was originally released under Creative Commons Attribution Non-Commercial.

## Glossary

ADHD: Attention-Deficit Hyperactivity Disorder

FC: Functional connectivity

dFC: Dynamic functional connectivity

WSBM: Weighted stochastic block model

PCA: Principal component analysis

SZ: Schizophrenia

BP: Bipolar disorder

EPI: Echo-planar imaging

SCID: Structured Clinical interview used for DSM disorders

BOLD: Blood oxygen level dependant

TS: Temporal stability

RSN: Resting state network

## Data Availability

All data produced in the present study are available upon reasonable request to the authors http://fcon_1000.projects.nitrc.org/indi/retro/cobre.html

## Acknowledgements

Data collection and sharing for this project (*Dataset 1)* was provided by UCLA Consortium for Neuropsychiatric Phenomics LA5c study. The public database was obtained via openfMRI (https://openfmri.org/dataset/ds000030/). The work was supported by the Consortium for Neuropsychiatric Phenomics (NIH roadmap for Medical Research grants UL1-DE019580, RL1MH083268, RL1MH083269, RL1DA024853, RL1MH083270, RL1LM009833, PL1MH083271 and PL1NS063410. The COBRE (The Centre for Biomedical Research Excellence) dataset *(Dataset 2)* was obtained through the International Neuroimaging Data-sharing initiative (http://fcon_1000.projects.nitrc.org/indi/retro/cobre.html). The imaging data and phenotypic information was collected and shared by the Mind Research Network and University of New Mexico funded by National institute of health centre of biomedical research excellence (COBRE) grant 1P20RR021938-01A2.

## Funding

AB acknowledges the generous support of the NBRC Flagship program BT/ MEDIII/ NBRC/ Flagship/ Program/ 2019: Comparative mapping of common mental disorders (CMD) over lifespan. We acknowledge the generous support of NBRC Core funds and the Computing facility.

## Author contributions

NS and AB conceptualized the study, NS undertook formal analysis, AB provided resources and tools and was involved in funding acquisition, NS wrote the first draft, both authors reviewed and edited the manuscript, AB supervised the work.

## Conflict of interest

The authors declare no conflict of interest.

## Supplementary Figures

**S 1.**
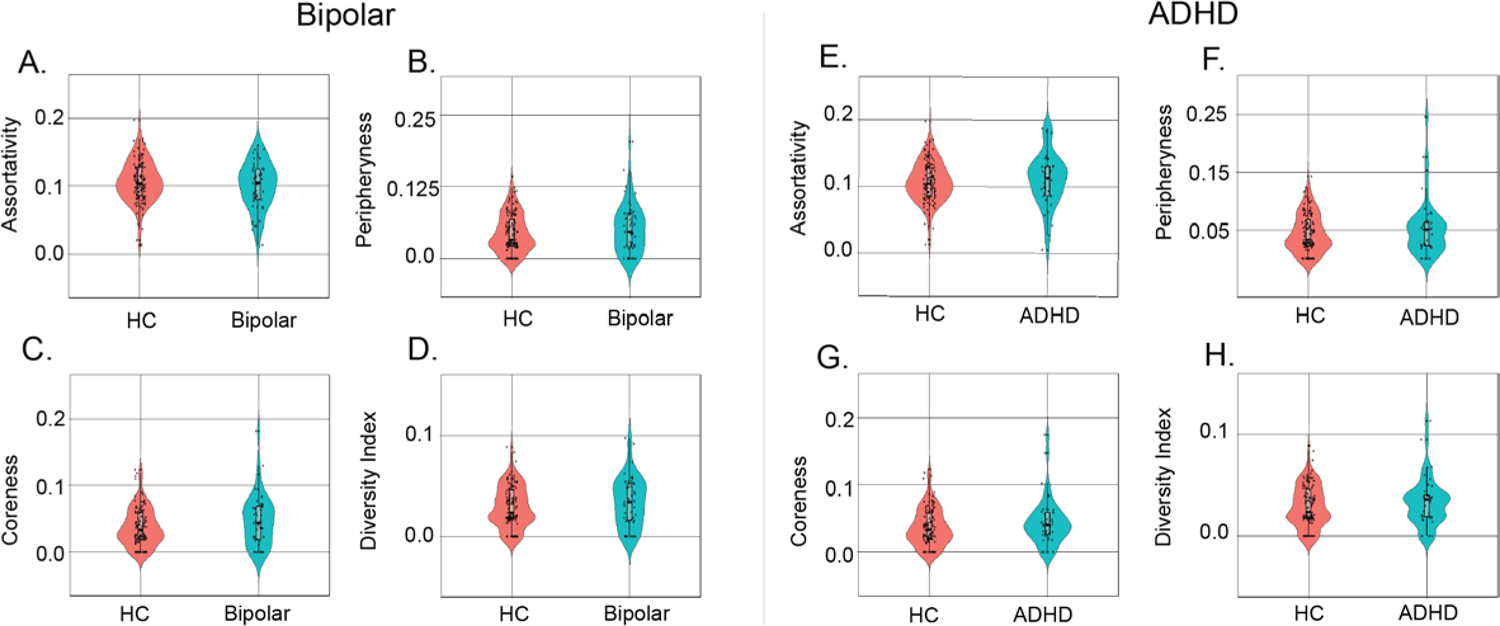
(A-D) Violin plots showing community interaction motifs (assortativity, coreness and peripheryness) and diversity index averaged across all communities per subject in participants with bipolar disorder (green) and healthy controls (orange) (E-F) Violin plots showing community interaction motifs (assortativity, coreness and peripheryness) and diversity index averaged across all communities per subject in participants with ADHD (green) and healthy controls (orange)

**S 2.**
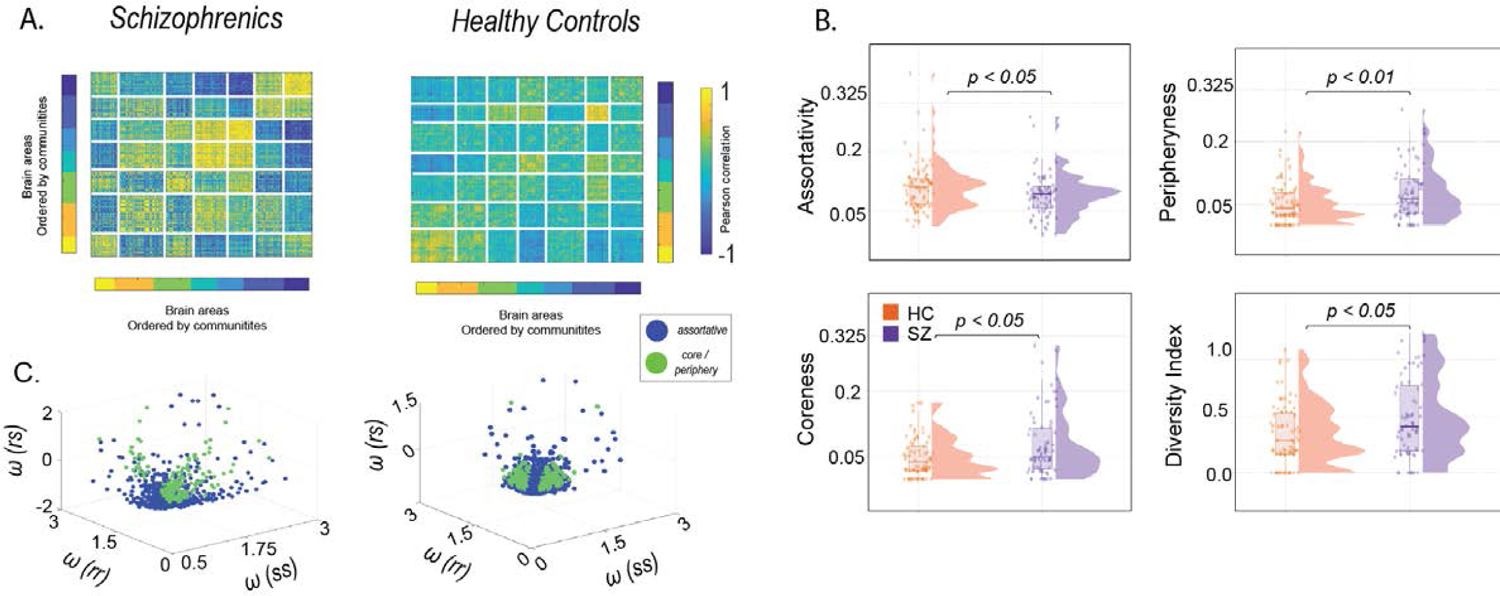
(A) Static functional connectivity matrices where each entry is the Pearson correlation between brain regions *n* and *p* ordered by communities (B) Each pair of communities, *r and s* are classified into one of three community motifs – assortative, coreness and peripheryness. Diversity index averaged across all brain regions per subject. The violin plots indicate, in schizophrenics communities are less assortative (C) Morphospace constructed by using all pairs of community interactions and are colour coded – blue (assortative community interactions) and green (core or peripheryness community interaction).

**S 3.**
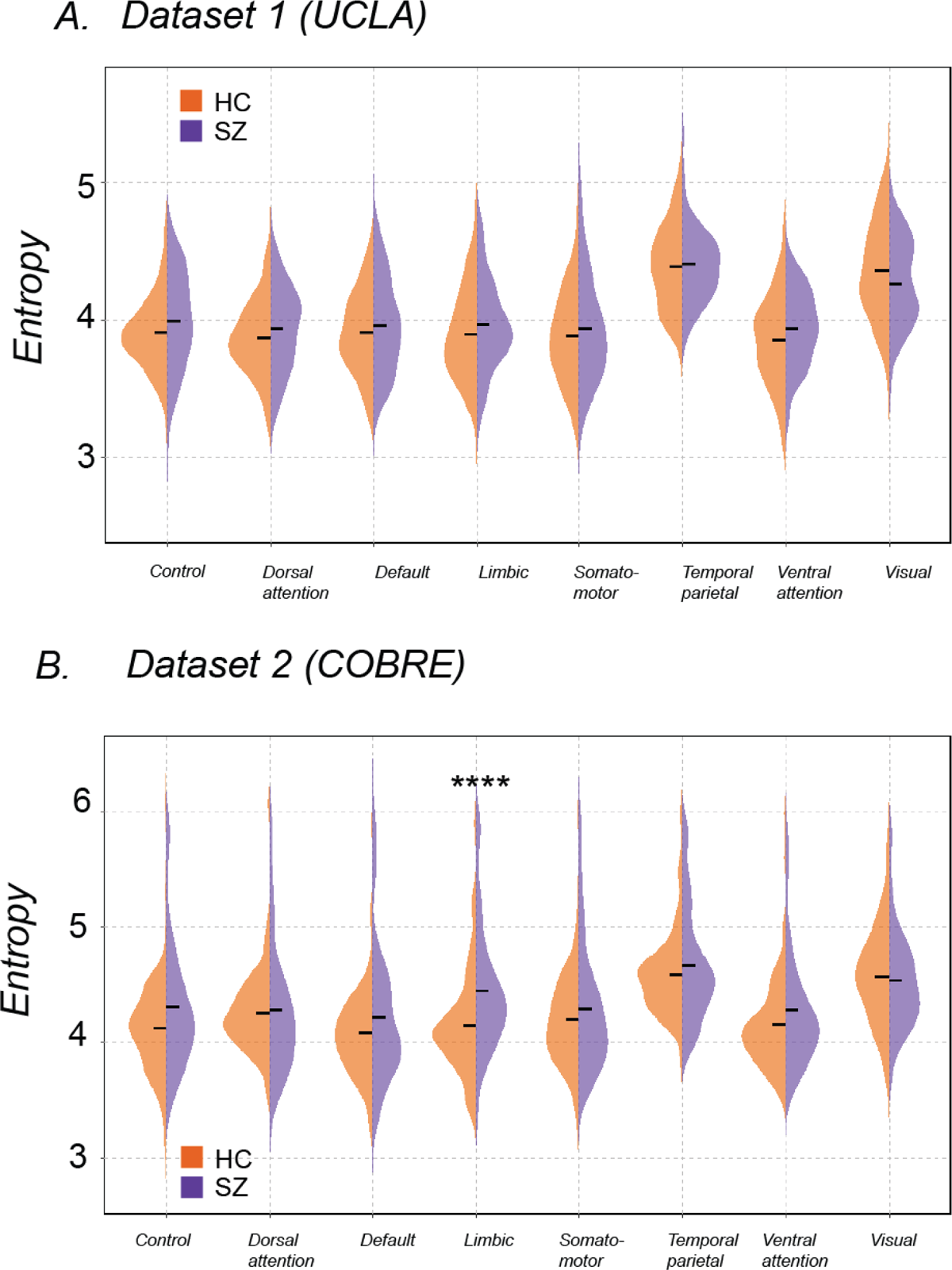
Quantifying temporal stability using entropy for the resting state networks defined in Schaeffer atlas in *Dataset 1* (A) and *Dataset 2* (B). Statistically significant differences (uncorrected) are indicated using * *(P≤ 0.05), ** (P≤ 0.01), ***(P≤ 0.001), ****(P≤ 0.0001),* ns (not significant).

**Table S1.** Demographics of the UCLA consortium for Neuropsychiatric phenomics LA5c dataset *(Dataset 1)* and COBRE dataset *(Dataset 2)* used in this study.

|  | Groups | N | Age (years) | Sex (% female) |
| --- | --- | --- | --- | --- |
| UCLA dataset<br>*(Dataset 1)* | Schizophrenia | 50 | 36.46 ± 8.87 | 24 % |
|  | Healthy Controls | 124 | 31.58 ± 8.80 | 44.90 % |
|  | Bipolar disorder | 49 | 35.28 ± 9.02 | 42.80 % |
|  | ADHD | 40 | 33.09 ± 10.7 | 51.16 % |
| COBRE Dataset<br>*(Dataset 2)* | Schizophrenia | 71 | 38.16 ± 13.8 | 19.71 % |
|  | Healthy Controls | 74 | 35.82 ± 11.5 | 31.08 % |

**Table S2.** Region wise significant assortativity differences in patients with schizophrenia, age and sex matched healthy controls.

| Brain regions<br>(ROI number) | Controls | Patient | P value |
| --- | --- | --- | --- |
| Peripheral visual (14) | 0.933333 | 0.886667 | 0.02618 |
| Peripheral visual (16) | 0.926667 | 0.9 | 0.03743 |
| Peripheral visual (217) | 0.926667 | 0.886667 | 0.03239 |
| Peripheral visual (222) | 0.94 | 0.906667 | 0.0004645 |
| Dorsal attention A (62) | 0.93 | 0.913333 | 0.04117 |
| Dorsal attention A (65) | 0.926667 | 0.926667 | 0.004987 |
| Dorsal attention A (69) | 0.943333 | 0.903333 | 0.02541 |
| Ventral attention/<br>Salient network (88) | 0.946667 | 0.916667 | 0.01481 |
| Ventral attention/(101)<br>Salient network | 0.933333 | 0.933333 | 0.01064 |
| Temporal parietal (395) | 0.913333 | 0.873333 | 0.007664 |

**Table S3.**
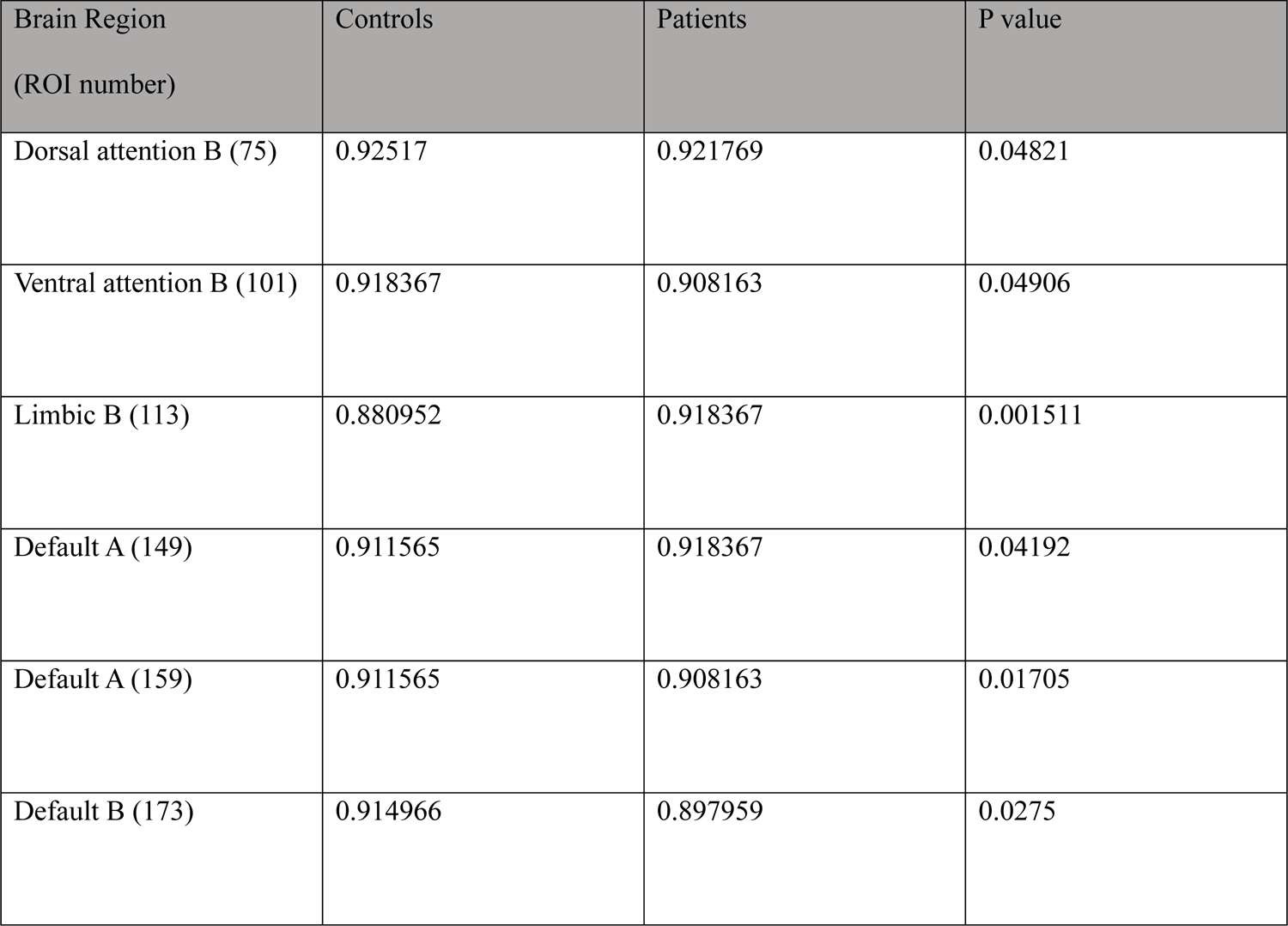
Region wise significant assortativity differences in patients with bipolar disorder, age and sex matched healthy controls.

**Table S4.** Region wise significant assortativity differences in patients with ADHD, age and sex matched healthy controls.

| Brain Regions<br>(ROI number) | Control | Patient | P value |
| --- | --- | --- | --- |
| Central visual (7) | 0.9 | 0.933333 | 0.02668 |
| Dorsal attention B (84) | 0.904167 | 0.870833 | 0.02501 |
| Limbic A (118) | 0.866667 | 0.866667 | 0.04766 |
| Control A (127) | 0.941667 | 0.9375 | 0.02998 |
| Control B (142) | 0.9 | 0.925 | 0.04677 |
| Default A (162) | 0.891667 | 0.858333 | 0.02264 |
| Default C (191) | 0.916667 | 0.883333 | 0.03971 |
| Default C (194) | 0.870833 | 0.85 | 0.04202 |
| Somatomotor A (227) | 0.929167 | 0.9 | 0.04864 |
| Limbic A (324) | 0.870833 | 0.875 | 0.04558 |
| Control A (331) | 0.9125 | 0.9 | 0.02631 |

## Notes

### Competing Interest Statement

The authors have declared no competing interest.

### Author Declarations

We downloaded resting state functional magnetic resonance imaging (fMRI) data from 285 participants who participated in the University of California Los Angeles (UCLA) Consortium for Neuropsychiatric Phenomics LA5c study. The public database was obtained via openfMRI (https://openfmri.org/dataset/ds000030/) A publicly available dataset from the centre for Biomedical Research Excellence (COBRE) was obtained obtained through the International Neuroimaging Data-sharing initiative (http://fcon_1000.projects.nitrc.org/indi/retro/cobre.html). This was originally released under Creative Commons Attribution Non-Commercial.

