## Supplementary document for "Dynamicity of brain network organization and their community architecture as characterizing features for classification of common mental disorders from the whole-brain connectome"

**Affiliation: 1 Cognitive Brain Dynamics Laboratory, National Brain Research Centre, NH 8, Manesar, Gurgaon 122052**

****

**Participants and Image Acquisition**

Dataset 1

We downloaded resting state functional magnetic resonance imaging (fMRI) data from 285 participants who participated in the University of California Los Angeles (UCLA) Consortium for Neuropsychiatric Phenomics LA5c study *(Dataset 1)* (Poldrack, et al., 2016) (Gorgolewski, Durnez , & Poldrack, 2017). The public database was obtained via openfMRI (https://openfmri.org/dataset/ds000030/) and includes 138 healthy controls (HC), 58 individuals diagnosed with schizophrenia (SZ), 40 with attention deficit hyperactivity disorder (ADHD) and 49 with bipolar disorder (BP). Data from 14 healthy controls and 8 participants with schizophrenia were removed during preprocessing (see below). For both healthy and patient groups, participants were men or women, of any racial group, whose primary language was either English or Spanish, who completed at least 8 years of formal education, no significant medical illness, had visual acuity 20/60 or better and urinalysis negative for drugs of abuse (Cocaine; Methamphetamine; Morphine; THC; and Benzodiazepines) (Poldrack, et al., 2016). In the healthy control group, participants were excluded if they had lifetime diagnosis of Schizophrenia, Bipolar I or II disorder, substance abuse/dependence or current major Depressive disorder, suicidality, anxiety disorder and ADHD. Healthy participants were also screened for threshold ADHD and they could not have had medication treatment for ADHD within the prior 12 months. Each of the patient groups (SZ, BP, ADHD) excluded anyone with one of these other diagnoses. Stable medications were permitted for the patients. Participants who were left-handed, pregnant or had other conditions (metal in the body) were excluded for MRI studies (Poldrack, et al., 2016). The resting state fMRI (rs-fMRI) data was acquired on 3T Siemens Trio scanner using echo planar imaging (EPI) sequence consisting of 152 volumes with the following parameters – slice thickness = 4mm, TR = 2s, TE = 30ms, flip angle = 90°, acquisition matrix = 64 X 64 sq.mm, voxel size = 3 X 3 X 4 $mm^3$. More details on acquisition parameters can be found in (Poldrack, et al., 2016).

Dataset 2

For the replication analysis, a publicly available dataset from the center for Biomedical Research Excellence (COBRE) was obtained (Calhoun, et al., 2012) (Bellec, 2016). The neuroimaging dataset *(Dataset 2)* included resting state functional MRI scans from 72 participants with schizophrenia and 74 healthy controls. All the subjects were screened and excluded if they had a history of mental retardation, a history of severe head trauma with more than 5 mins of loss of consciousness, history of substance abuse or dependence within the last 12 months. Diagnostic information was collected using the Structured Clinical interview used for DSM disorders (SCID). The eyes open rs-fMRI data was collected with echo-planar imaging (EPI sequence) consisting of 150 volumes, scan duration of 5 mins with a repetition time (TR) = 2s, echo time = 29 ms, acquisition matrix = 64 X 64 sq.mm, flip angle = 75 ° and voxel size = 3 X 3 X 4 $mm^3$. A detailed description of acquisition parameters can be found in (Bellec, 2016).

**Data preprocessing**

The rs-fMRI images were pre-processed using the CONN toolbox (McGovern Institute for Brain Research, MIT, USA) in MATLAB (The MathWorks). The default CONN preprocessing pipeline (defaultMNI) was employed, consisting of functional realignment and unwarp, slice-time correction, outlier identification, direct segmentation and normalization, and functional smoothing. In the first step, the fMRI data were unwarped and realigned using SPM12 *realign & unwarp* procedure (Andersson, Hutton, Ashburner, Turner, & Friston , 2001) where all scans are co-registered to a reference image using a least squares approach, resampled using b-spline interpolation to correct for motion and magnetic susceptibility interactions. Temporal misalignment between different slices of the functional data was corrected using SPM splice-timing correction (STC) procedure (Sladky , et al., 2011). Potential outlier scans were identified using ART. Functional and anatomical scans were normalized to standard MNI space, segmented into grey matter, white matter and cerebro-spinal fluid (CSF) tissue classes, and resampled to 2mm isotropic voxels following a direct normalization procedure using SPM unified segmentation and normalization algorithm (Calhoun, et al., 2017). Lastly, functional data is smoothed using spatial convolution with a Gaussian kernel of 8 mm full-width half maximum (FWHM). In addition, functional data were denoised using a standard denoising procedure (Castanon, 2020), followed by bandpass frequency filtering of BOLD time series between 0.01 Hz and 0.1 Hz. In *Dataset 1*, an inspection of fMRI data for each subject resulted in the exclusion of 14 healthy controls and 8 schizophrenics whose data did not include – 1) all 152 functional volumes 2) T1 w structural images. A detailed overview of the preprocessing pipeline can be found at https://web.conn-toolbox.org/fmri-methods/preprocessing-pepeline .

**Data Analysis**

Model-based community detection using the weighted stochastic block model (WSBM) on static FC

The weighted stochastic block model (WSBM) is a generative model for learning community structure, which places each of the *n* nodes (brain areas) in the adjacency matrix *A* into one of *k* communities or “blocks” (Aicher , Jacobs , & Clauset, 2015). Nodes in the same community are stochastically equivalent, indicating their equivalent roles in generating the network’s structure. In its classic form stochastic block model (SBM), assumes an unweighted network, and the probability of edge existence is learned for each block. The weighted stochastic block model *(WSBM)* is a generalization of SBM that can learn from both the presence and weight of the edges. Specifically, WSBM models each weighted edge $A_{ij}$ as a draw from a parametric family distribution, whose parameters $\mu$ and $\sigma$ depend only on block memberships of connecting nodes $i$ and $j$ (Aicher , Jacobs , & Clauset, 2015) (Tooley , Bassett, & Mackey, 2022).

In the SBM, the network's adjacency matrix $A$ contains binary values for edge existence, $A_{ij}$ $\epsilon$ {0,1}, $k$ denotes the fixed number of blocks or communities and vector $z$ contains the group label for each node $Z_i \in \{1, 2, \ldots\ldots k\}$. The SBM assigns an edge existence parameter to each edge bundle $\theta_{k\acute{k}}$. Assuming the placement of the edges are independent of one another, the likelihood function of SBM for $A_{ij}$ can be written as:

$$\Pr(A \mid z, \theta) = \prod_{ij} \theta_{Z_i Z_j}^{A_{ij}} \; (1 - \theta_{z_i z_j})^{1-A_{ij}} \tag{9}$$

Which can be rewritten as,

$$\Pr(A \mid z, \theta) = \prod_{ij} \exp(A_{ij}.\log\left(\frac{\theta_{z_i z_j}}{1-\theta_{z_i z_j}}\right) + \log(1 - \theta_{z_i z_j})) \tag{10}$$

The community structure of WSBM retains the stochastic equivalence principle of SBM. In case of WSBM instead of edge-existence probabilities, each edge bundle is now parameterised by a mean and variance i.e., $\theta_{z_i z_j} = (\mu_{z_i z_j}, {\sigma^2}_{z_i z_j})$. The likelihood function would be,

$$\Pr(A \mid z, \mu, \sigma^2) = \prod_{ij} \mathrm{N}\left(A_{ij} \middle| \mu_{z_i z_j}, {\sigma^2}_{z_i z_j}\right) = \prod_{ij} \exp(A_{ij}.\log\left(\frac{\mu_{z_i z_j}}{{\sigma^2}_{z_i z_j}}\right) - A^2.\frac{1}{2{\sigma^2}_{z_i z_j}} - 1.\frac{{\mu^2}_{z_i z_j}}{{\sigma^2}_{z_i z_j}} \tag{11}$$

Where $\mu \in R^{k\,X\,k}$ and $\sigma^2 \in R^{k\,X\,k}$ are model parameters, $A_{ij}$ is the adjacency matrix, $\mu_{z_i z_j}$ $and$ ${\sigma^2}_{z_i z_j}$ parameterize the weights of edges between community $z_i$ and $z_j$, $\Pr(A \mid z, \mu, \sigma^2)$ denotes the probability of generating the network $A$ given the parameters.
